# Sinus and Anterior Skull Base Surgery during the COVID-19 pandemic: Systematic review, Synthesis and YO-IFOS position

**DOI:** 10.1101/2020.05.01.20087304

**Authors:** Thomas Radulesco, Jerome R. Lechien, Leigh J Sowerby, Sven Saussez, Carlos Chiesa-Estomba, Zoukaa Sargi, Philippe Lavigne, Christian Calvo-Henriquez, Chwee Ming Lim, Napadon Tangjaturonrasme, Patravoot Vatanasapt, Dehgani-Mobaraki Puya, Nicolas Fakhry, Tareck Ayad, Justin Michel

## Abstract

**Purpose:** The COVID-19 pandemic has caused significant confusion about healthcare providers’ and patients’ pandemic-specific risks related to surgery. The aim of this systematic review is to summarize recommendations for sinus and anterior skull base surgery during the COVID-19 pandemic.

**Methods:** PubMed/MEDLINE, Google Scholar, Scopus and Embase were searched by two independent otolaryngologists from the Young Otolaryngologists of IFOS (YO-IFOS) for studies dealing with sinus and skull base surgery during COVID-19 pandemic. The review also included unpublished guidelines edited by Otolaryngology-Head and Neck Surgery or Neurosurgery societies. Perioperative factors were investigated including surgical indications, preoperative testing of patients, practical management in operating rooms, technical aspects of surgery and postoperative management. The literature review was performed according to PRISMA guidelines. The criteria for considering studies or guidelines for the review were based on the population, intervention, comparison, outcome, timing and setting (PICOTS) framework.

**Results:** 15 international publications met inclusion criteria. Five references were guidelines from national societies. All guidelines recommended postponing elective surgeries. An algorithm is proposed that classifies endonasal surgical procedures into three groups based on the risk of postponing surgery. Patients’ COVID-19 status should be preoperatively assessed. Highest level of personal protective equipment (PPE) is recommended, and the use of high-speed powered devices should be avoided. Face-to-face postoperative visits must be limited.

**Conclusions:** Sinus and skull base surgeries are high-risk procedures due to potential aerosolization of SARS-CoV-2 virus. Protection of health care workers by decreasing exposure and optimizing use of PPE is essential with sinus and anterior skull base surgery.

## Introduction

Since its emergence in December 2019 in Wuhan, China, severe acute respiratory syndrome coronavirus 2 (SARS-CoV-2) has rapidly spread worldwide.[1] The pandemic has significantly impacted the management of all patients, in all medical fields, regardless of COVID-19 status. In many countries, the number of elective surgeries has been substantially reduced to redeploy healthcare workers and to preserve personal protective equipment (PPE), medication supply and hospital equipment for the care of COVID-19 patients [2].

An critical issue concerns the risk of infection for health care providers [3]. In Wuhan, 40 of the 138 first patients hospitalized were health care providers, and the same issue was reported in Italy.[4, 5] It has been shown that severe acute respiratory syndrome coronavirus 2 (SARS-CoV-2) is characterized by high viral loads in upper airways [6]. As such, Otolaryngologist – Head and Neck surgeons and surrounding staff are especially vulnerable to viral transmission during clinical encounters, in-office procedures and surgery [7, 8]. Among the common ENT surgeries, tracheostomy, endolaryngeal procedures and sinus and skull base surgeries are still considered as high-risk procedures [7]. There are currently only a few published recommendations specifically addressing sinus and anterior skull base surgery. Most elective surgeries are being postponed, but practitioners need recommendations to continue to manage urgent cases [9]. Moreover, the planning for the resumption of surgical activity after the first peak of the pandemic in the next months must be anticipated and planned at this time.

The aim of this systematic review was to summarize national recommendations or publications related to sinus and anterior skull base surgery during the COVID-19 pandemic, focusing on surgical indications, preoperative testing of patients, practical management in operating rooms, technical aspects of surgery and postoperative management.

## Methods

The study was conducted by the COVID-19 Task Force of the Young Otolaryngologists of the International Federation of Oto-rhino-laryngological Societies (YO-IFOS), which includes European, Asian, North and South American, Oceanian and Middle-East and African otolaryngologists. The literature review was performed according to PRISMA guidelines [10] (**Figure 1**). Two independent authors conducted the PubMed/MEDLINE, Google Scholar, Scopus, Embase and MedRxiv search for identifying papers published between January 2020 and April 2020. Titles and abstracts were reviewed titles and abstracts to screen out non-relevant articles and the working group reviewed the full text of remaining articles/guidelines. The following keywords were used for the search strategy: “COVID-19” OR “SARS CoV-2” AND “ENT” OR “sinus surgery” OR “transnasal” OR “nasal” OR “endoscopic” OR “neurosurgery” OR “skull base” OR “guidelines” OR “consensus”. This review also included unpublished guidelines and recommendations edited by different otolaryngological or neurosurgical societies and institutions across the world, by contacting societies directly.

**Figure 1.**
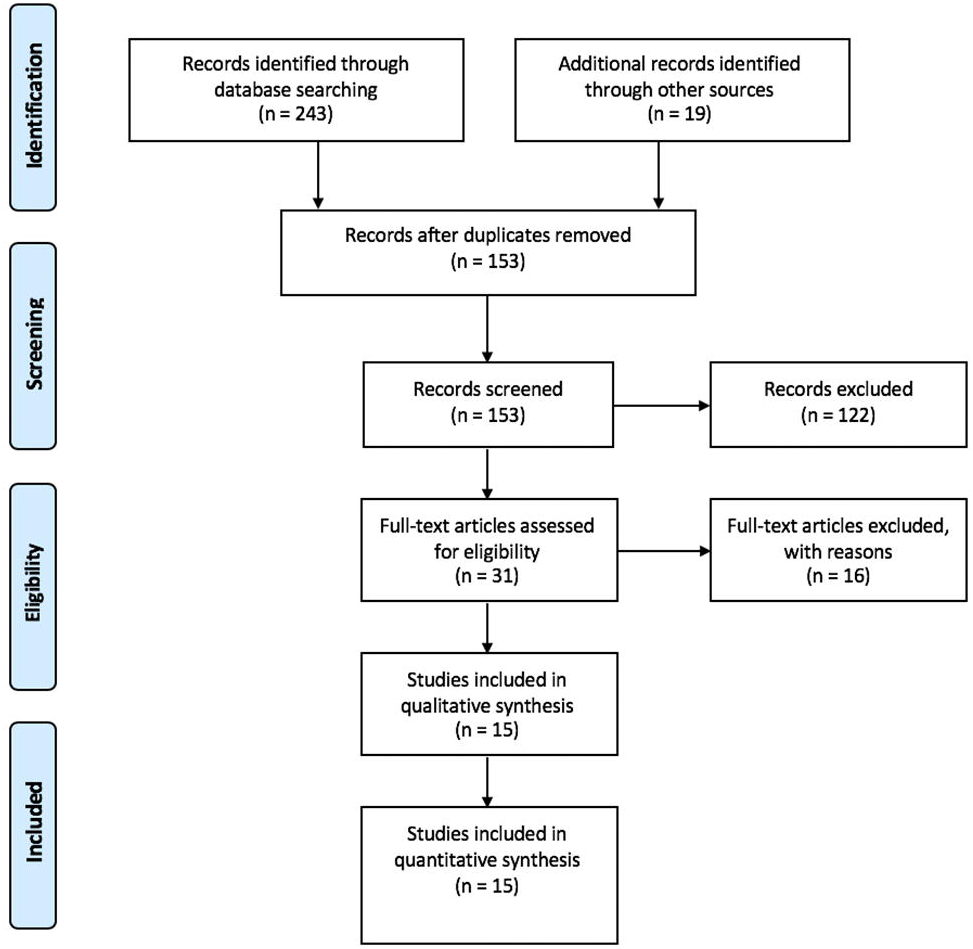
PRISMA diagram

The criteria for considering studies or guidelines for the review were based on the population, intervention, comparison, outcome, timing and setting (PICOTS) framework [11] [12].

### Participants and inclusion criteria

Authors had to provide substantial information about management of patients undergoing sinus or skull base surgery. No language restriction was applied to the search strategy. Cadaveric studies were not considered in the present review.

### Intervention

All publications and/or guidelines related to sinus or anterior skull base surgery and COVID-19 were included in this review.

### Comparison and Outcome

The primary outcome was to produce a set of recommendations to improve sinus and skull base surgery safety. We compared surgical indications, preoperative testing of patient’s strategy, general management in operating rooms (OR), Personal Protective Equipment (PPE), technical aspects of surgery and postoperative management.

### Timing

All publications referred to COVID-19 pandemic period. Guidelines and publications from January 2020 were included. All searches were completed on April 18^th^, 2020.

### Setting

Publications from community, private and tertiary care university hospitals were included.

## Results

The initial search strategy found 153 results of which 15 met inclusion criteria. References originated from the United Kingdom [13], France [14], Italy [15][16][17] United States [7, 18–21], China [22–24], Australia [25] and one was an international consensus statement [26] (**Table 1**). Five references were guidelines from national ENT or neurosurgical societies [13, 17–19, 25]. Publication dates ranged from February 2, 2020 to April 17, 2020.

**Table 1.**
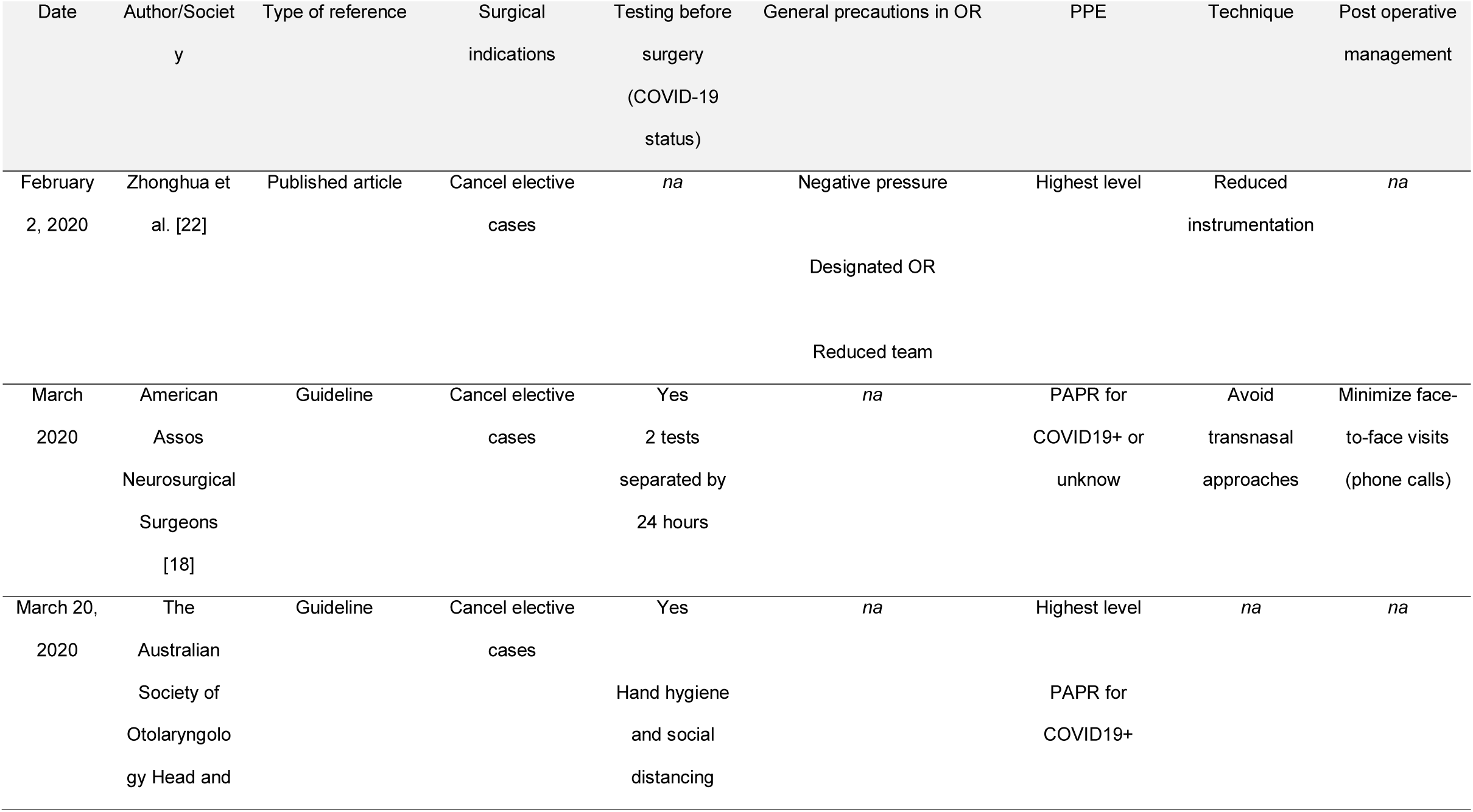
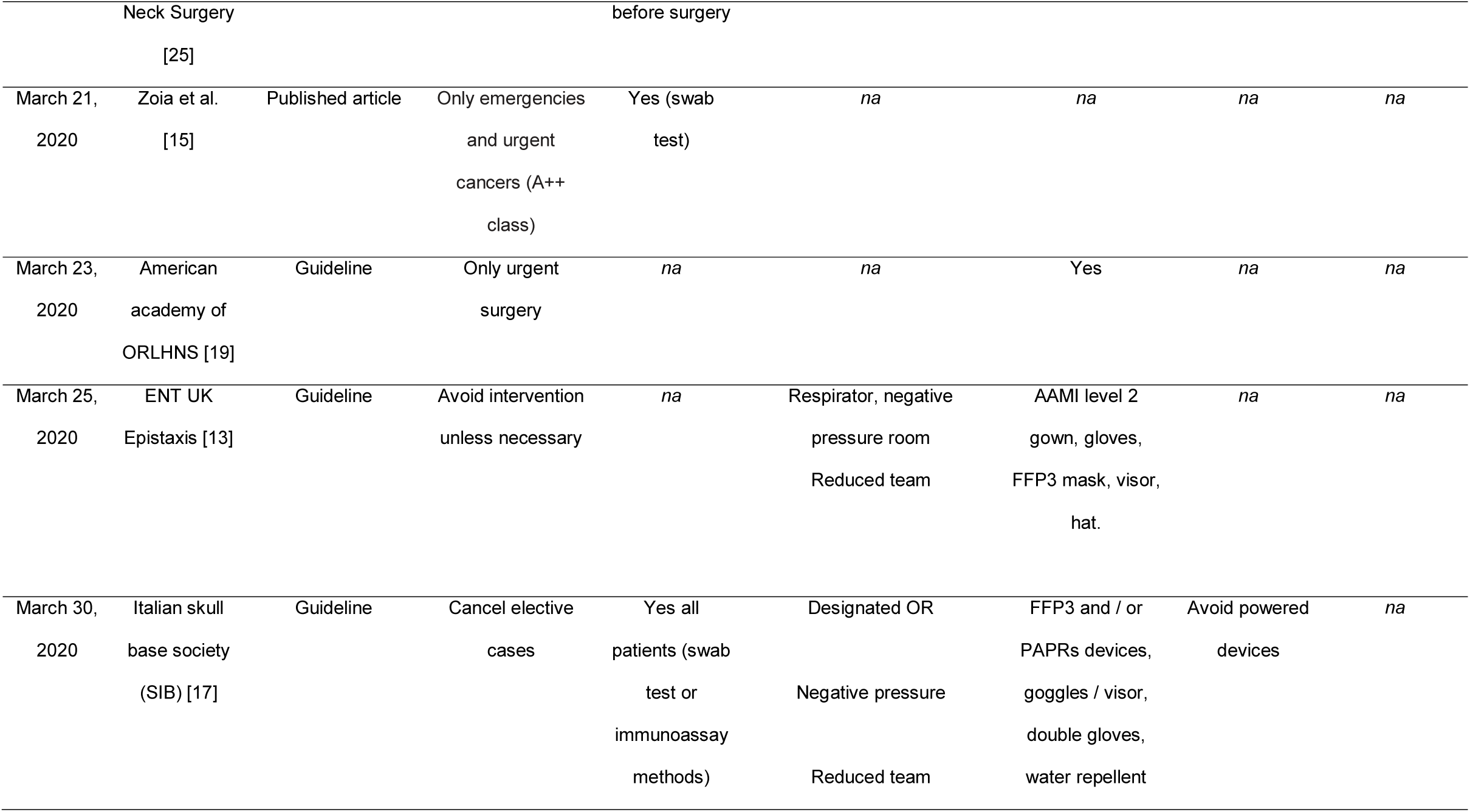
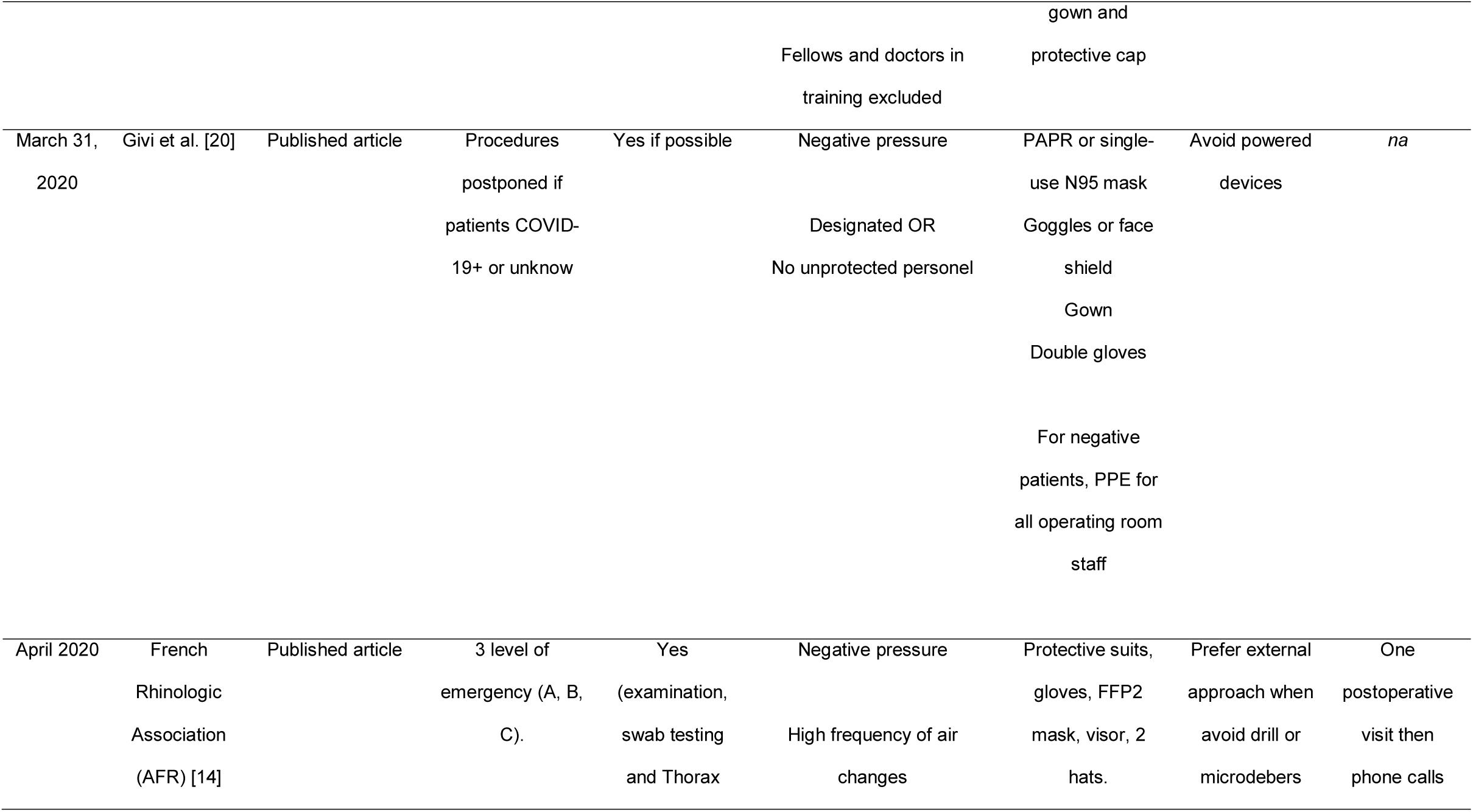
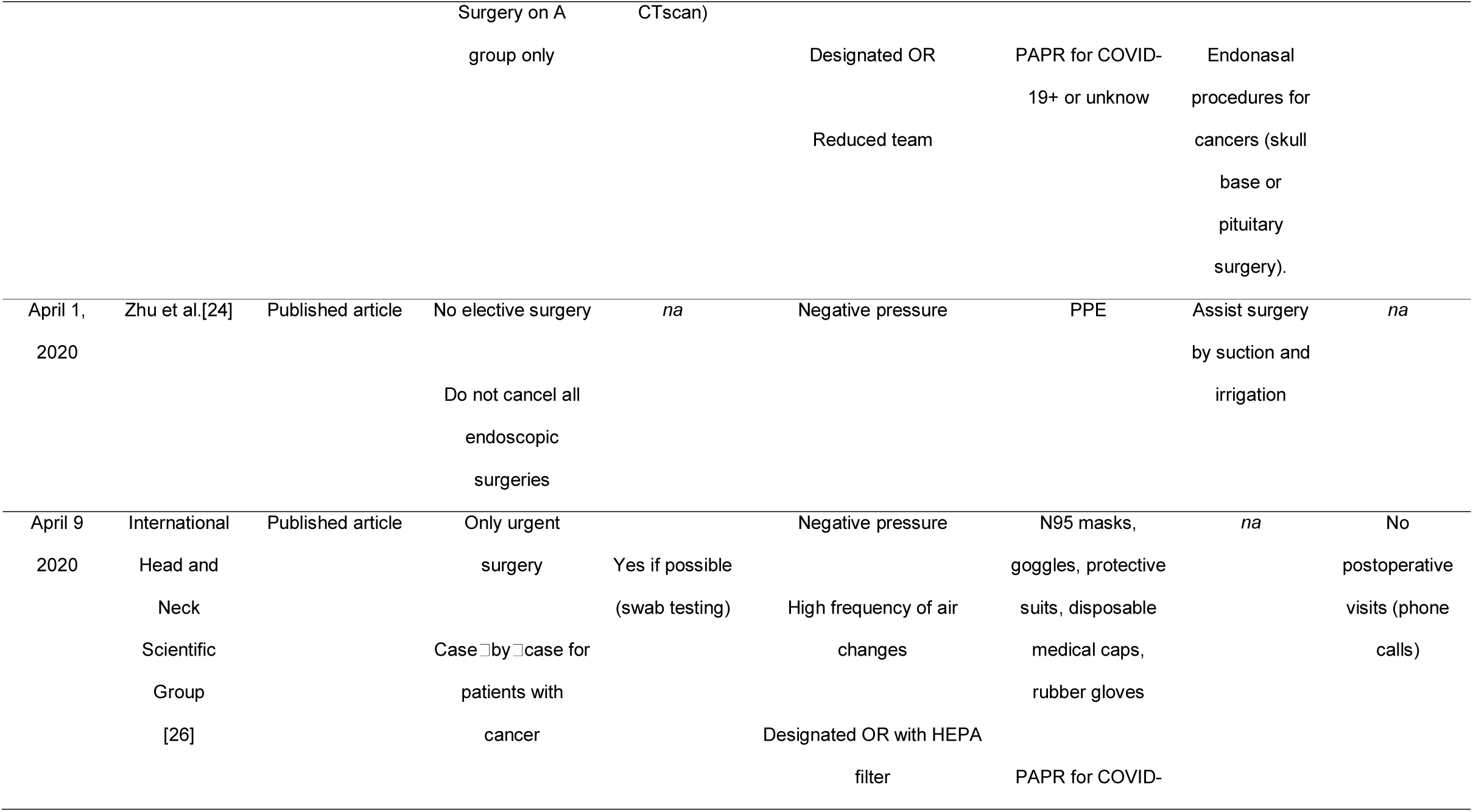
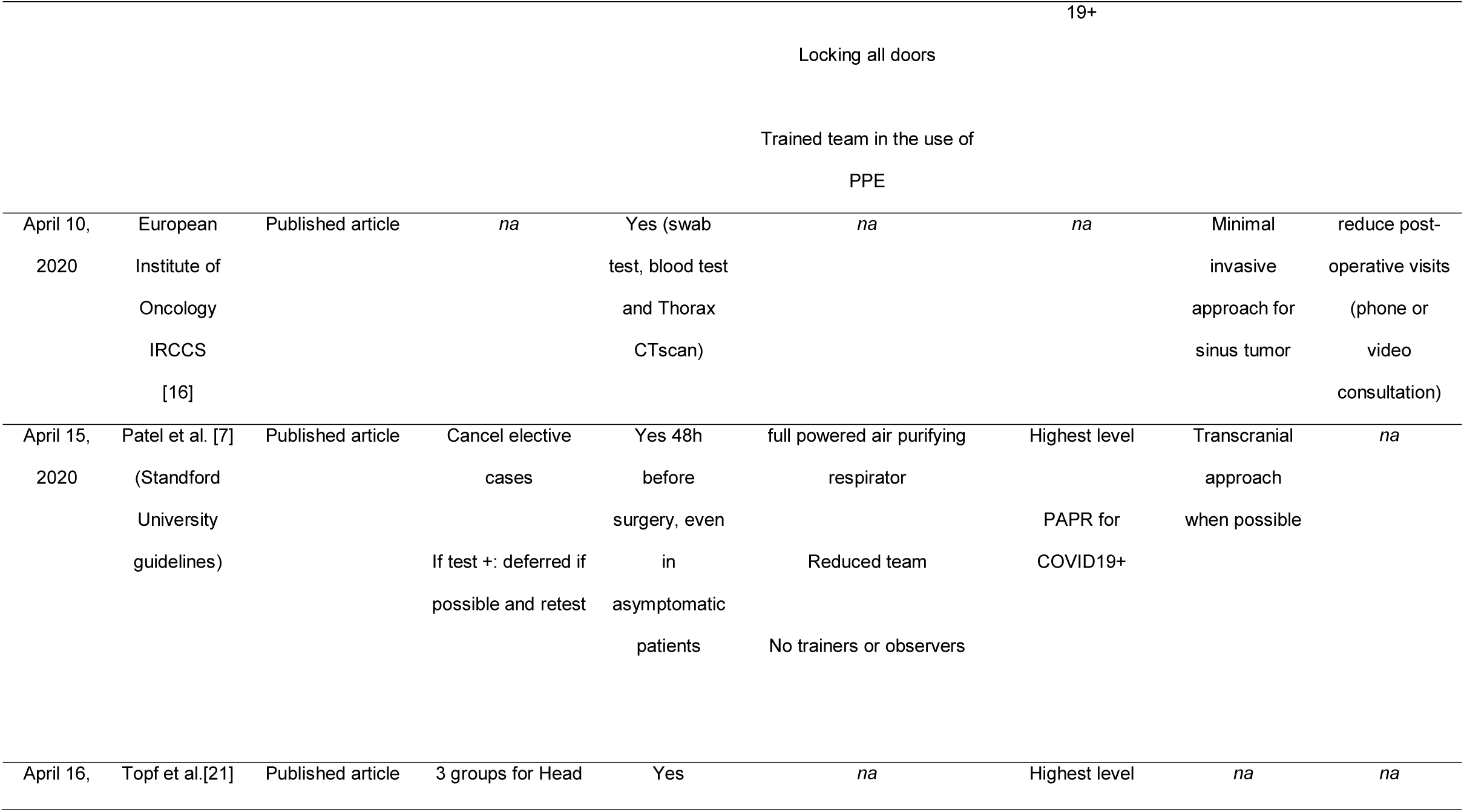
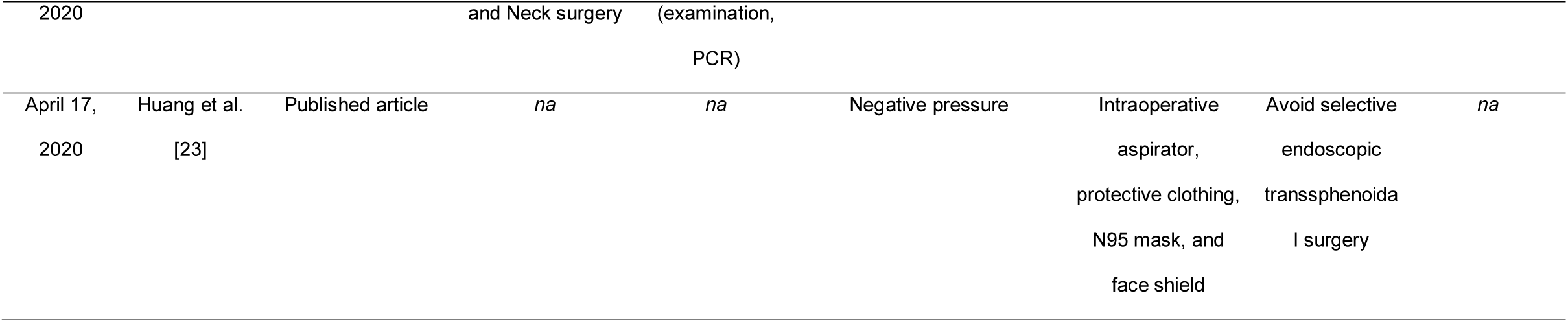
Title: Published articles and international guidelines included in the review. Legend: Published articles and international guidelines concerning sinus and skull base surgery during COVID-19 pandemic. Abbreviations: OR = Operative Room; HEPA = High-Efficiency Particulate Air; PAPR = Powered Air Purifying Respirators; PPE = Personal Protective Equipment

### Surgical indications

The postponing of elective sinus and skull base surgeries was recommended in almost all guidelines [7, 13–15, 17–22, 24–26]. Only urgent (and case by case consideration for patients with cancer) procedures were suggested to be performed during the pandemic. The French Rhinology Association (Association Française de Rhinologie, AFR) proposed an algorithm classifying surgical procedures into three groups: - A: requiring immediate treatment - B: requiring treatment within a maximum of 4 weeks and - C: non-urgent procedures. The updated algorithm was validated by YO-IFOS working group (**Table 2**). In France, only group A procedures could be performed during the pandemic. Group B procedures would then be prioritized once group A procedures were performed.

**Table 2.**
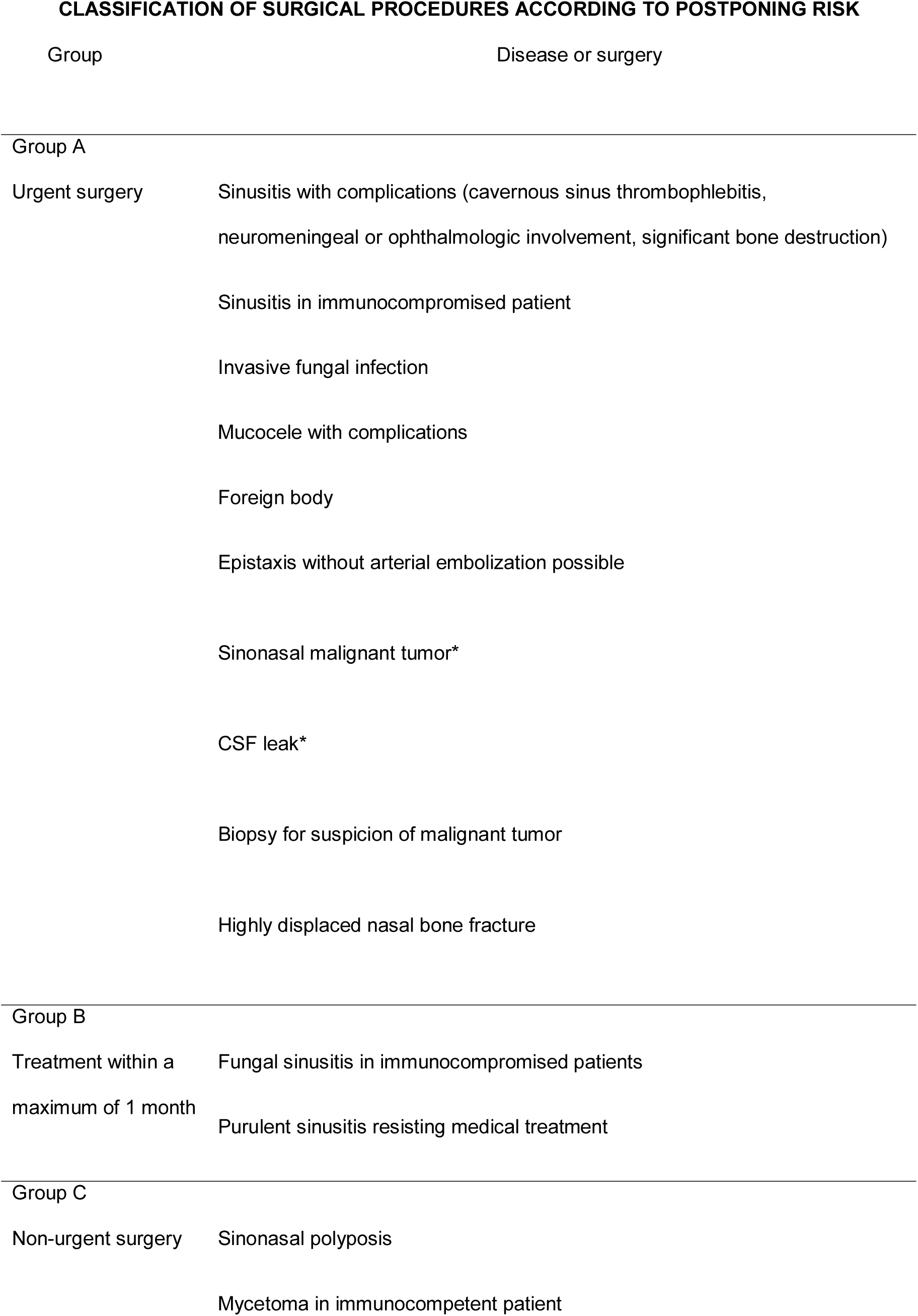
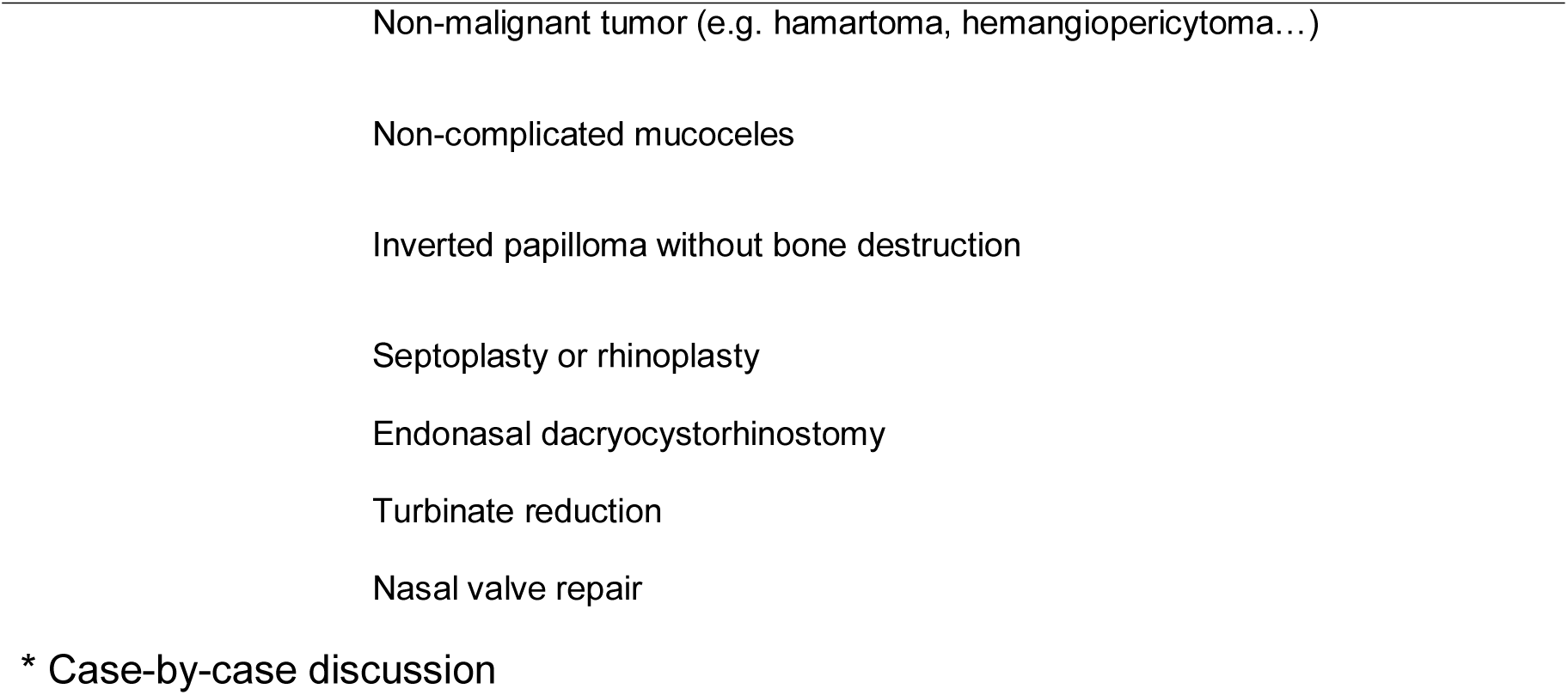
Title: Classification of surgical procedures according to postponing risk. Legend: Classification or sinus or skull base surgery according to postponing risk derived from AFR proposition [14].

CLASSIFICATION OF SURGICAL PROCEDURES ACCORDING TO POSTPONING RISK
| Group | Disease or surgery |
| --- | --- |
| Group A |  |
| Urgent surgery | Sinusitis with complications (cavernous sinus thrombophlebitis, neuromeningeal or ophthalmologic involvement, significant bone destruction) |
|  | Sinusitis in immunocompromised patient |
|  | Invasive fungal infection |
|  | Mucocele with complications |
|  | Foreign body |
|  | Epistaxis without arterial embolization possible |
|  | Sinonasal malignant tumor* |
|  | CSF leak* |
|  | Biopsy for suspicion of malignant tumor |
|  | Highly displaced nasal bone fracture |
| Group B |  |
| Treatment within a maximum of 1 month | Fungal sinusitis in immunocompromised patients |
|  | Purulent sinusitis resisting medical treatment |
| Group C |  |
| Non-urgent surgery | Sinonasal polyposis |
|  | Mycetoma in immunocompetent patient |

\* Case-by-case discussion

### Preoperative assessment of patients’ COVID-19 status (RT-PCR, chest computed tomography, examination)

Radulesco recommended to contacting patients by phone 48 hours before surgery to ensure that the patient had no symptoms suggestive of COVID-19 [14]. However, several patients may present with mild symptoms, and up to 80% of cases have no symptoms at all.[27] Patients must be aware of hand hygiene and social distancing before surgery [25]. Most of authors recommended preoperative testing of patients using one or two nasopharyngeal swab tests [7, 14–18, 20, 21, 25, 26]. However, with false negative rates of RT-PCR approaching 30%, it has been suggested that patients requiring high-risk surgery (e.g. sinus or skull base surgery using drills) be treated as COVID-19 positive [28]. False negative rates could be decreased by repeating a second COVID-19 swab at least 24 hours apart and as close as possible to the date of surgery. Coupling nasopharygeal swab tests with chest computed tomography (CT-scan) for characteristic COVID-19 findings has been suggested as another method for overcoming false negative rates [14, 16][29]. The authors recognize, however, that resource limitations may not allow for RT-PCR or chest CT-scan to be carried out. The Italian Society of Skull base surgery (SIB) has also tried immunoassay methods with good results [17].

### General precautions in the Operating Room (OR)

Negative pressure rooms and designated COVID-19 ORs are the most commonly cited general measures[7, 13, 14, 17, 20, 22–24, 26]. High frequency (25 per hour) of air exchanges and the use of HEPA (High-Efficiency Particulate Air) filters can also help with cleaning the air [26]. The OR team must also be reduced to decrease potential healthcare worker exposure, and proper training in the use of PPE is of great importance [7, 13, 14, 17, 22, 26]. Patel *et al*. and SIB recommend that no trainees or observers be allowed in OR to save PPE in resource-constrained situations [7, 17].

### Personal Protective Equipment (PPE)

The use of highest level of PPE was recommended by 12 authors for all COVID-19+ or unknown patients, including FFP2 (N95) or FFP3 (N99) masks, goggles or face shield, fluid-resistant surgical gown, double gloves and a head covering [7, 13, 14, 17–26]. Suggestions have been made for consideration of use of personal air-purifying respirators (PAPRs) as they provide a higher level of protection than an N95 mask and are also reusable [7, 14, 17, 18]. Lastly, it is critical to properly perform donning and doffing of PPE to avoid contamination during and after the case.

### Technical specifics for sinus and anterior skull base surgery

Based on the study of Workman *et al*., three authors agreed that the use of high-speed drills should be avoided [14, 17, 20, 30]. Indeed, the use of drills was shown to generate a significant aerosol contamination of particles measuring >20um [30]. Regarding surgical approach, four guidelines recommended external approaches when amenable to avoid the use of endonasal endoscopes or powered devices (e.g. Caldwell-Luc, transcranial approaches) [7, 14, 18, 23].

Regarding anterior skull base surgery, the endonasal approach remains superior because external approaches could also increase aerosolization through the use of high speed instrumentation [14, 17]. Some authors propose the use of additional intraoperative aspirators (smoke evacuators) or a suction tube in the oral cavity to create a negative pressure environment inside the patient’s nasal or pharyngeal cavity [23, 24]. This method has not been evaluated in the literature for efficacy.

### Postoperative visits

To curb the spread of the COVID-19 pandemic, 4 authors recommended limiting postoperative visits [14, 16, 18, 26].

The YO-IFOS working group recommendations are summarized in **Table 3**.

**Table 3.**
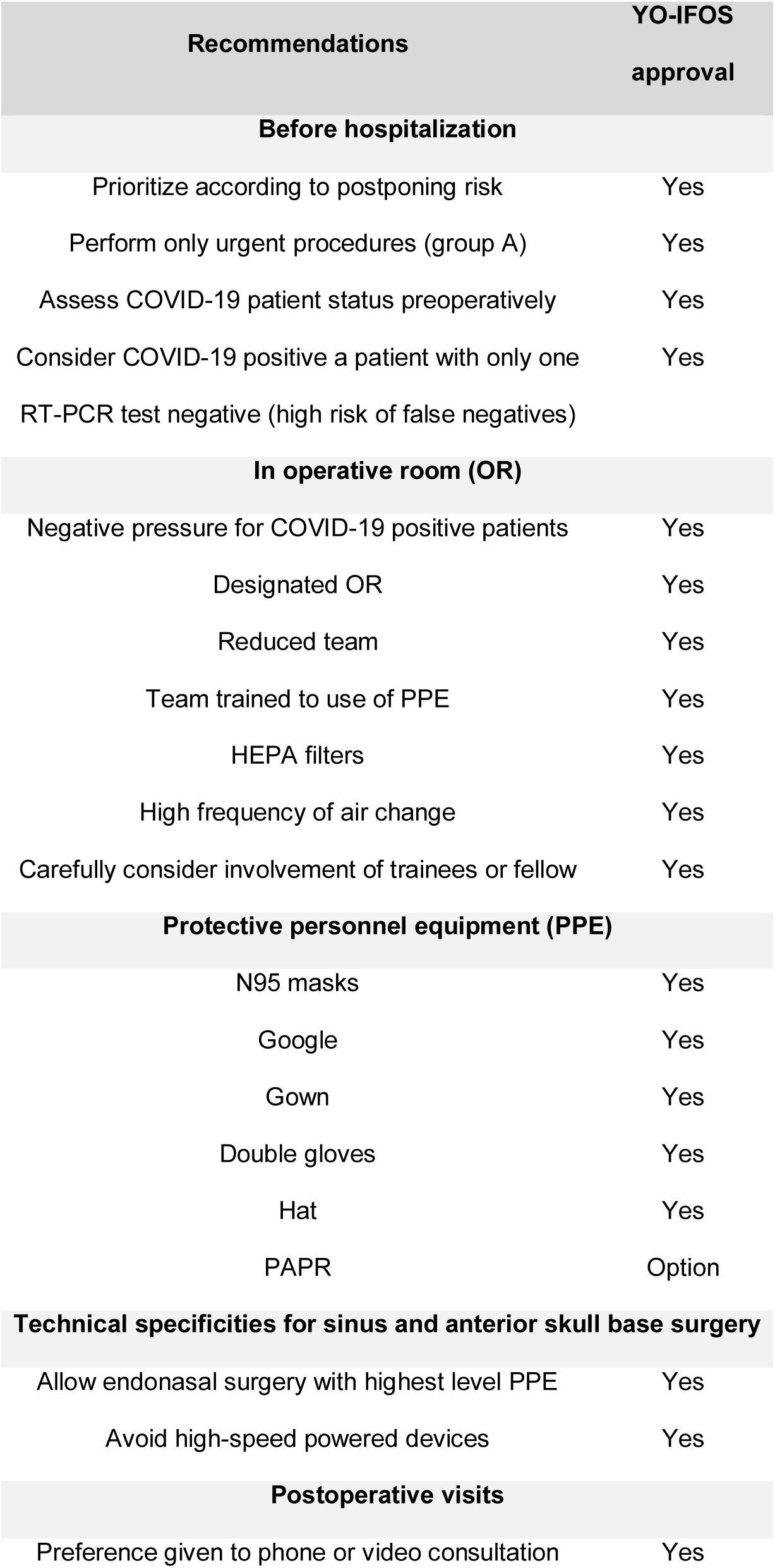
Title: Summary of YO-IFOS recommendation Legend: Summary of YO-IFOS recommendation regarding sinus and anterior skull base surgery. Abbreviations: OR = Operative Room; HEPA = High-Efficiency Particulate Air; PAPR = Powered Air Purifying Respirators; PPE = Personal Protective Equipment

| **Recommendations** | **YO-IFOS approval** |
| --- | --- |
| **Before hospitalization** |  |
| Prioritize according to postponing risk | Yes |
| Perform only urgent procedures (group A) | Yes |
| Assess COVID-19 patient status preoperatively | Yes |
| Consider COVID-19 positive a patient with only one RT-PCR test negative (high risk of false negatives) | Yes |
| **In operative room (OR)** |  |
| Negative pressure for COVID-19 positive patients | Yes |
| Designated OR | Yes |
| Reduced team | Yes |
| Team trained to use of PPE | Yes |
| HEPA filters | Yes |
| High frequency of air change | Yes |
| Carefully consider involvement of trainees or fellow | Yes |
| **Protective personnel equipment (PPE)** |  |
| N95 masks | Yes |
| Goggles | Yes |
| Gown | Yes |
| Double gloves | Yes |
| Hat | Yes |
| PAPR | Option |
| **Technical specificities for sinus and anterior skull base surgery** |  |
| Allow endonasal surgery with highest level PPE | Yes |
| Avoid high-speed powered devices | Yes |
| **Postoperative visits** |  |
| Preference given to phone or video consultation | Yes |

## Discussion

The previous statements reflect the practice of sinus and anterior skull base surgery management during the COVID-19 pandemic and the YO-IFOS position on this. These data may change depending on local public health aspects of the pandemic and its impact on the healthcare system over time. The availability of masks and other protective gear continues to be a challenge in many parts of the world. The usefulness of PPE is no longer debatable and protection of health care workers by reducing exposure is a key-point regarding sinus and skull base surgery. Improving access to PPE and safely extending the duration of use allows for better protection of surgeons and surrounding staff. The use of powered air purifying respirators (PAPR) was recommended in 6 publications when performing high-risk procedures, (e.g. using drills) but there was little data on the infection risk compared with other methods, either alone or in combination with the use of a mask [7, 14, 17, 18, 20, 25, 26]. These recommendations are based on Patel’s article [7], reporting a communication with surgeons in Wuhan. Surgeons directly involved in these cases, however – as reported by Huang – clarified misunderstandings around the surgical case and warn against unnecessary anxiety created toward endonasal endoscopic procedures based on an anecdotal statement [23]. No convincing evidence exists to show that there is an increased possibility of infection from endoscopic anterior skull base surgery without PAPRs at this time. The availability of this protective equipment is often limited and should be discussed on a case-by-case basis. Furthermore, the CDC (Centers for Control Disease and Prevention) and many individual hospitals do not support their use in the OR.[31] Otherwise, standard PPE use remains the best protective measure in response to COVID-19+ or unknown patients requiring surgical intervention.

The alternative to reducing surgical activity would be to systematically and accurately establish every patient’s COVID-19 status prior to surgery. The low number of RT-PCR tests available and their reliability are major pitfalls in applying such recommendations for now. Based on the local stage of the pandemic, the availability of screening tests, PPE and healthcare system capacity remain the most important factors to take into account when applying these general recommendations. Only two authors consider CT-scan for COVID-19 status assessment [14, 16]; a recent study found that chest CT-scan sensitivity was much greater than that of RT-PCR (98% vs 71%, respectively, p<.001) [32]. The relatively low number of group A procedures and the inherent high viral load within the nasal cavity are an argument to use both CT and RT-PCR to assess COVID-19 status before sinonasal surgery. To date, available recommendations regarding surgical indications only relate to cancer surgery, although other diseases can be considered similarly urgent [15, 21]. **Table 2** is a non-exhaustive list of surgical indications for which surgeries may proceed at the discretion of the respective hospital and surgeon.

The development of telephone or video consultation is an alternative to face-to-face general practice consultations and offers an appropriate option in certain settings [33]. Phone or computer-assisted visits should be offered whenever possible.

The main limitation of these recommendations is the disparity that may exist between centers regarding the availability of screening tests or protective equipment, influencing authors’ point-of-view. In addition, most of the references were written in an emergency crisis involving a lower quality level of evidence.

### Perspectives

Once the peak of the pandemic has passed, all these items must be reevaluated to encompass evolving evidence. Concerning surgical indications, most of patients of group B patients should be operated on within weeks. This will considerably increase the number of surgical procedures. In this period, COVID-19 will still be a concern. Great attention should be paid to continue the protective measures. Preoperative testing strategy will depend on test availability, population incidence and health policy. As Otolaryngologists, it is our duty to remind administrators and policy makers that Otolaryngology remains one of the most exposed and vulnerable specialties when it comes to the risk of COVID-19 transmission. We should continue pushing for pre-operative testing to be able to safely continue to take care of patients. Serology testing or vaccination will likely result in major changes when they become available [34]. The development of tools for video consultations or meetings should be considered as an opportunity to limit one-on-one contact [35]. If carefully employed, It will likely lead to long-term modification of our practices with a concurrent increase in patient satisfaction [36].

Future health policy may integrate city, state/province or country incidence rates. If there are no active cases in a region, perhaps having a patient self-quarantine for 7-10 days prior to surgery would be sufficient to effectively rule out SARS-COV-2.

## Conclusion

Sinus and skull base surgery are high-risk procedures due to potential aerosolization of viral particles. Surgical indications must be prioritized according to patient’s risk and the ability of health care providers to have adequate protection. Practitioners, where possible, should use video or phone consultations to limit the risk of transmission.

## Data Availability

all data referred to in the manuscript and note links are available

